# Assessing the acceptability of a malaria prevention intervention in infant by healthcare providers: the contribution of structural equation modeling (SEM)

**DOI:** 10.1101/2025.09.01.25334879

**Authors:** Natacha Revollon, Shino Arikawa, Elodie Richard, Arnold Sadio, Rodion Konu, Bandana Bhatta, Martin Kouame Tchankoni, Koku Delanyo Dzoka, Diane Fifonsi Gbeasor-Komlanvi, Cristina Enguita Fernandez, Francisco Saute, Mohamed Samai, Valérie Briand, Bernard Atchrimi, Didier Koumavi Ekouevi, Joanna Orne-Gliemann, MULTIPLY project consortium

**Author notes:** corresponding author (NR).

## Abstract

**Introduction:** Despite evidence of its efficacy and cost-effectiveness, and being recommended by WHO since 2010, the implementation of preventive malaria prophylaxis for infants (strategy now called perennial malaria chemoprevention) has remained limited in sub-Saharan Africa. The MULTIPLY project aimed to pilot, in rural Togo, this strategy extended in the 2nd year of life and integrated within facility-based and outreach vaccination services. Our aim was to conduct an in-depth study of the acceptability of perennial malaria chemoprevention prior to its implementation, in order to inform future programmatic deployment of this strategy.

**Methods:** A cross-sectional Knowledge, Attitudes and Practices questionnaire was self-administered pre-implementation of perennial malaria chemoprevention in Haho district, among 231 health care providers. Three acceptability components, derived from the Theoretical Framework of Acceptability, were investigated: ease of administration, perceived effectiveness and necessity of the strategy. Structural equation modeling was used to identify the complex relationships between acceptability components, feasibility factors, knowledge, attitudes and practices.

**Results:** The overall pre-implementation acceptability of perennial malaria chemoprevention was high. Administration of perennial malaria chemoprevention was perceived as being all the easier if it was integrated into a setting where feasibility criteria, such as the presence of human resources (β= 0.18; p<0.01), were met. Perceived effectiveness of the strategy was associated with the knowledge of the strategy (β= 0.20; p<0.01) and perception of drug effectiveness (β= 0.48; p<0.01). Perceived effectiveness of malaria control programs (β= 0.23; p<0.01) and low perception of the effectiveness of insecticide-treated bednets (β= -0.10; p=0.04) was associated with the perceived need for perennial malaria chemoprevention. Health care providers’ seniority was associated, directly or indirectly, with all three dimensions of the acceptability of perennial malaria chemoprevention.

**Conclusion:** The use of structural equation modeling allowed a comprehensive and nuanced quantitative assessment of pre-intervention acceptability of perennial malaria chemoprevention, highlighting the importance of considering feasibility factors such as access to and availability of human and material resources and attitudes towards malaria prevention when promoting the adoption of this childhood preventive strategy among health care providers in a rural district in Togo.

## Introduction

Since 2010, the World Health Organization (WHO) has recommended intermittent preventive treatment of malaria in infants (IPTi) with sulfadoxine-pyrimethamine (SP) in areas of moderate to high transmission^1^ [1,2]. IPTi is a pragmatic intervention, integrated within the Expanded Program on Immunization (EPI)[2,3]. IPTi was shown to reduce malaria morbidity [4], is well tolerated [5–7] and cost-effective [8]. However by 2020, only Sierra Leone was routinely implementing IPTi in sub-Saharan Africa [9]. The main challenges to IPTi implementation reported at community level were geographical accessibility, mistrust of the healthcare system [10] and low vaccination coverage [11]. At the health care system level, in spite of health-care providers’s acceptance of IPTi [10], the lack of equipment, training and high staff turnover have limited the effective implementation of the strategy [11,12].

Since June 2022, the WHO has re-endorsed the IPTi strategy, extending it to the 2nd year of a child’s life and renaming it perennial malaria chemoprevention (PMC) [13]. PMC may present additional challenges to implementers, as compared to IPTi, including the decline in vaccination coverage over time [14]. Documenting the acceptability and feasibility of PMC [15] may contribute to improve its effective implementation and help save lives. In the context of the MULTIPLY project piloting PMC in 3 sub-saharan African countries, an initial PMC readiness assessment of the health district of Haho in Togo [16], guided among others by the Theoretical Framework of Acceptability [17], showed that PMC was perceived as effective and *a priori* easy to administer, although some providers did not understand its value for children who are not sick. This work also helped to identify potential obstacles to the future implementation of PMC in Togo, notably insufficient staff in EPI services, or lack equipment and access to health centers. Although this initial assessment of readiness and acceptability helped to adapt the implementation of PMC locally, it highlighted also that the acceptability of a health intervention is a complex and multifactorial concept [17]; several questions emerged: Why do some providers perceive PMC as effective but not as necessary to prevent malaria in children? Do providers’ knowledge, attitudes and practices have an influence on the acceptability of PMC? Is the perceived feasibility of PMC associated with its acceptability?

In this paper, we aimed to assess the interactions between the different aspects and components of PMC acceptability in Togo, and the factors influencing this acceptability, in order to identify the most relevant levers to promote the effective implementation of PMC at lower levels of care in sub-Saharan Africa.

## Materials and methods

### Context

Malaria is endemic in Togo: in 2022, the prevalence of malaria infection was estimated at 32% among children aged 10 to 23 months, in the Plateaux region [18]. The MULTIPLY project aimed to pilot a PMC strategy in the rural district of Haho, consisting in the administration of 4 doses of SP during the first two years of life, via the EPI platform, both facility-based and during outreach services [19] [16]. A comprehensive mixed methods assessment of the MULTIPLY-PMC strategy were carried out, in order to assess its effectiveness in real-life conditions, acceptability and feasibility, cost-effectiveness and impact [19].

### Study design

We conducted a cross-sectional Knowledge, Attitudes and Practices (KAP) survey in November 2021, prior to the implementation of the MULTIPLY-PMC strategy in Haho district of Health Care Workers (HCWs) and Community Health Workers (CHWs) [16].

### Study population and sampling

The study population consisted of 188 HCWs and 525 CHWs practicing in all 27 health facilities in Haho district in 2021. Mixed sampling was carried out: exhaustive for all 188 HCWs and random stratified by health facility for CHWs (10% of the workforce per facility).

### Data collection methods and tools

A KAP questionnaire, designed by a multidisciplinary team based on pre-existing malaria KAP questionnaires and adapted to include PMC-relevant questions, aimed to document: respondents’ socio-demographic data, health facility characteristics, knowledge, attitudes and practices regrading malaria, preventive measures in children, and PMC.

Questionnaires were distributed by disctrict health authorities, and self-administered in paper format, between November 1 and 30, 2021. All participants were asked to provide their informed consent in writing before participating. Anonymized data were entered by project staff into the REDCap database, and were available from April 1, 2022. Data confidentiality and anonymity were preserved throughout the collection, analysis, and interpretation process.

### Available variables and coding

Three KAP variables were designed to specifically measure the pre-intervention acceptability of PMC, each one corresponding to a specific component of the TFA [17]: perceived efficacy (variable *Effective*); perceived burden, estimated by the perceived ease of administering treatment (variable *Easy to administer*); and intervention coherence, estimated by the perceived need to administer treatment to a child who is not ill (variable *Necessary*). Five variables documented attitudes towards immunization and malaria prevention strategies. The perceived feasibility of PMC was documented by four *proxi* variables on the perception that facility staff and equipment was sufficient to deliver immunization services. Acceptability, attitudes, and feasibility variables had 4 response modalities coded from 1 to 4 (from *Strongly agree* to Strongly *disagree)*.

Knowledge variables had multiple response options, documenting understanding of malaria causes, symptoms, and means of prevention. A global knowledge score (maximum 23 points) was computed as follows: each response was allocated 1 point if correct and zero if not; questions on basic knowledge (transmission by mosquito, for example) were worth 2 points if correct.

Selected practices variables consisted in systematic information for parents coded from 1 to 3 and correct registration coded from 1 to 5.

### Data analysis

#### Descriptive analysis

A descriptive analysis of all socio-demographic, acceptability, attitudes, feasibility and practices variables was carried out and presented in the form of means and standard deviations for continuous variables, and in the form of numbers and frequencies for categorical variables.

#### Structural equation modeling

We conducted a post hoc exploratory analysis (i.e., not planned and designated prior to data collection) to assess the links and interactions between the components of acceptability, using structural equation modeling (SEM). Widely used in the field of social and behavioral sciences since the 1970s [20], the SEM method allows to identify complex relationships between dependent variables (here, the 3 components of acceptability), with independent quantitative variables belonging to different domains (here, feasibility factors, knowledge, attitudes and practices). Briefly, SEM analysis includes: i) defining a conceptual model and specifying hypothetical relationships between these different variables, based on the literature; ii) checking whether these hypothetical relationships are confirmed by our study dataset (direct effects) and iii) identifying whether there are correlations between the variables included in the model (indirect effects) [20,21].

The conceptual model (Fig 1) represents the hypotheses of the variables likely to influence the pre-intervention acceptability of the MULTIPLY-CPP strategy; the hypotheses (Table 1) were formulated on the basis of the literature.

**Fig 1.**
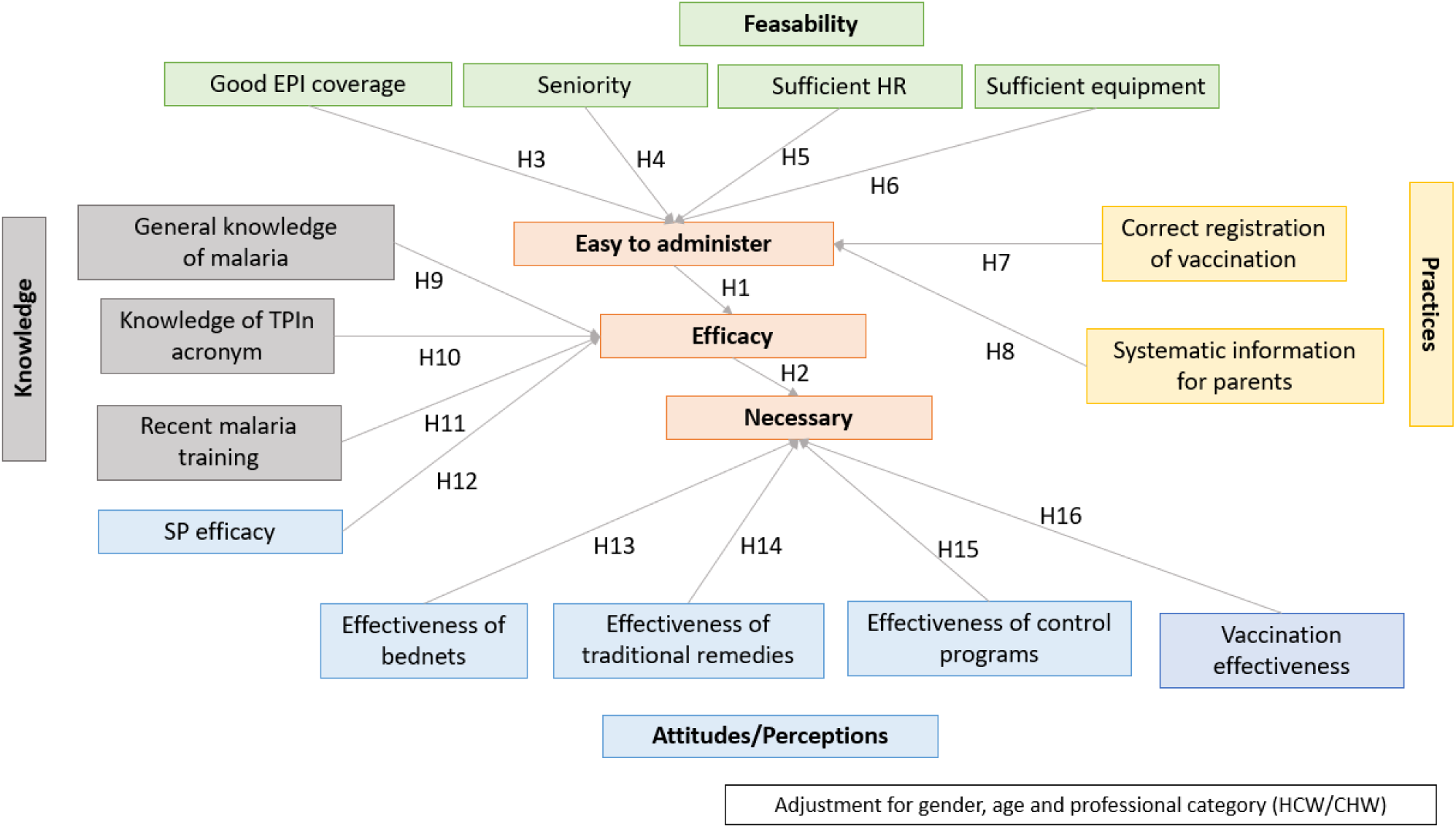
Conceptual model of the acceptability of PMC, MULTIPLY project, Haho district, Togo, November 2021

**Table 1.**
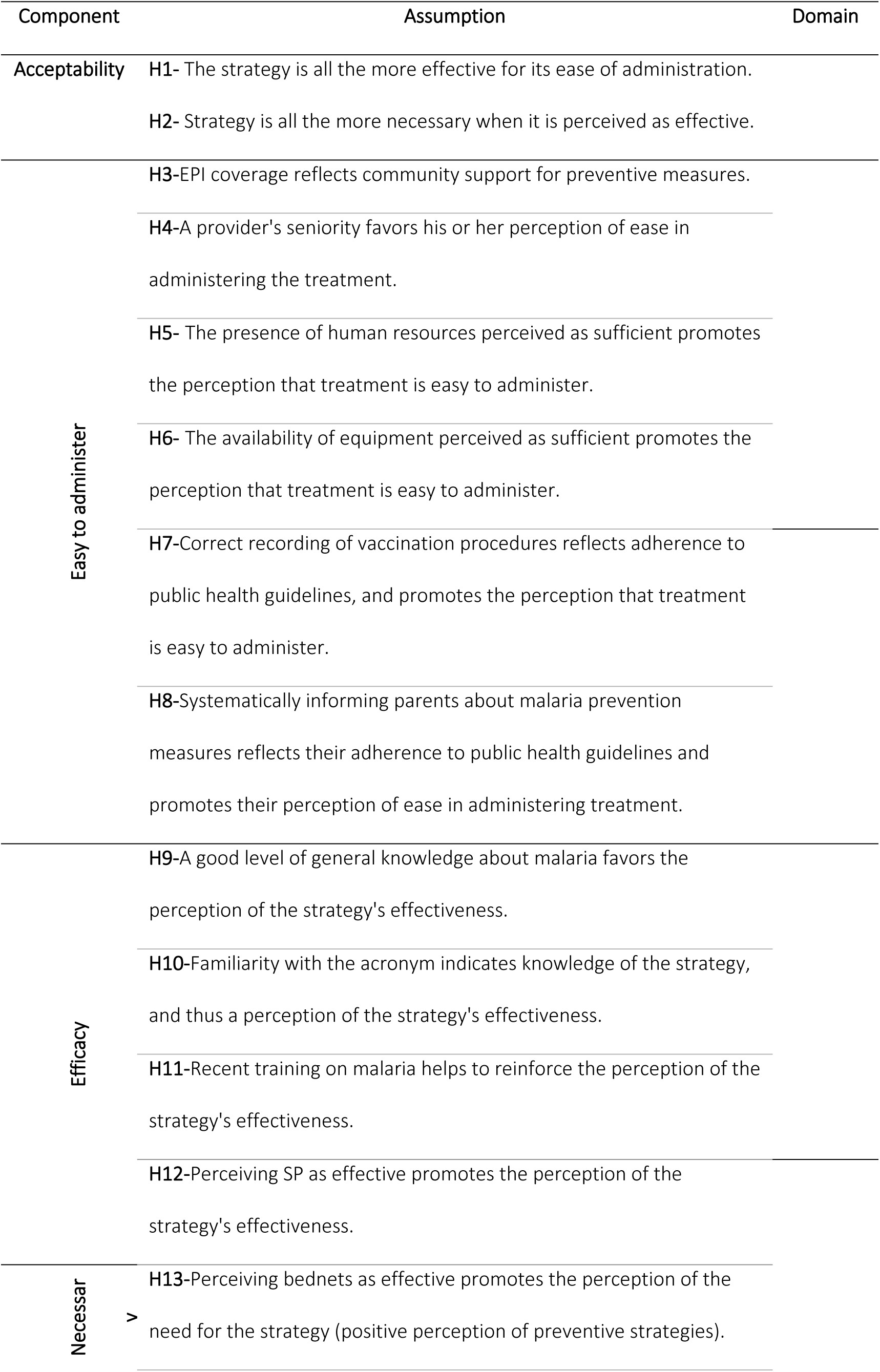
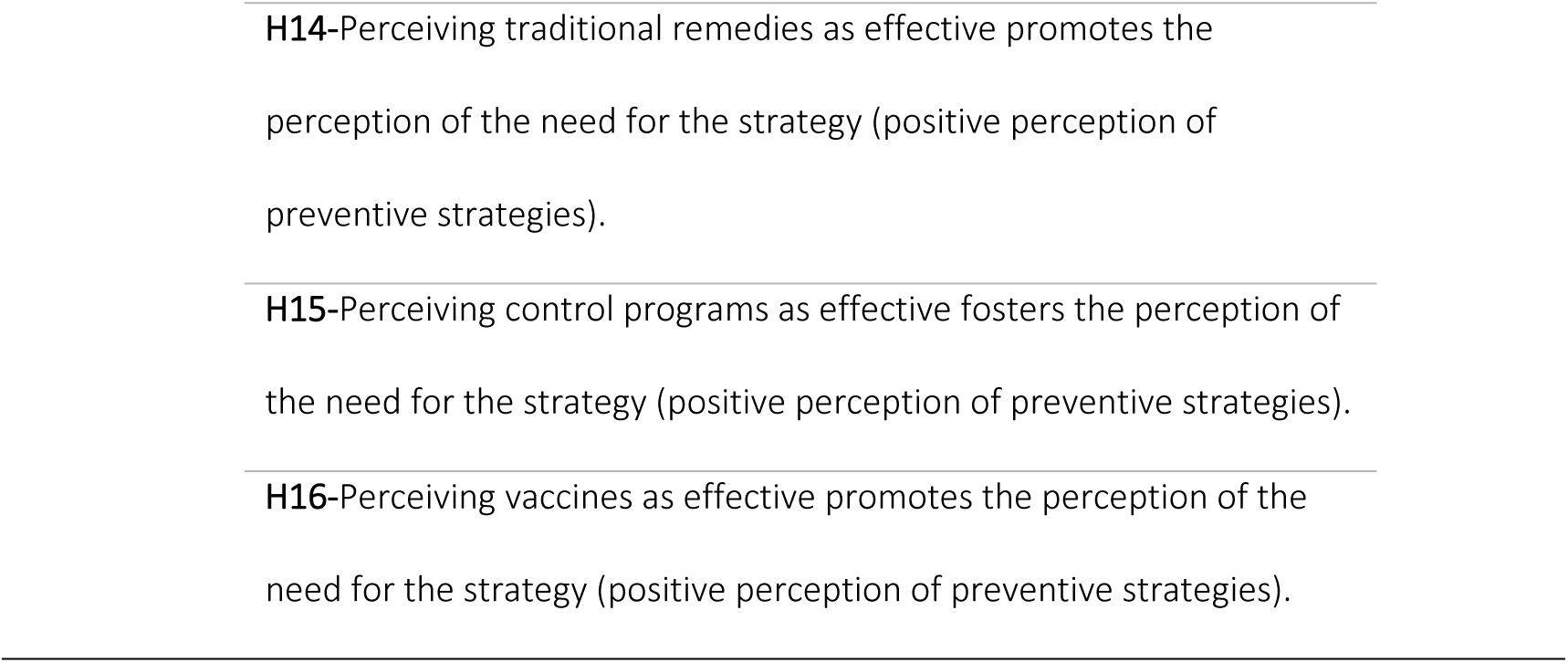
List of assumptions made in the conceptual model

We hypothesized that the perceived ease of PMC administration would be supported by: perceptions of community adherence to EPI schedules [9,11,22–27], the provider’s seniority [9,11], or the perceptions of sufficient human resources [11,25] and equipment within the health facility [9,11,25] (H3 to H6, Table 1). Practices reflecting adherence to public health guidelines could also facilitate the implementation of a new strategy recommended by the public authorities [22] (H7 and H8 Table 1). Another hypothesis is that the perceived effectiveness of PMC could be influenced by health care providers’ levels of knowledge and training [9,11,22,23], and their perceived effectiveness of SP [11,23,28] (H9 to H12 Table 1). Further, the perception of the need to administer malaria chemoprevention could be influenced (positively or negatively) by attitudes towards traditional preventive remedies [22,24] (H14 Table 1). Finally, we hypothesised (no available literature yet) that 1) health care providers’ perception of the need to administer PMC could be influenced by the perceived efficacy of other preventive strategies such as the use of bednets, malaria control programs and vaccination (considering PMC is integrated within the EPI platform) (H13, 15 and 16 Table 1) and 2) the 3 components of PMC acceptability could influence each other – PMC would be perceived as effective if also perceived as easy to administer, and perceived as needed if also perceived as effective (H1 and H2, Table 1).

Age, gender, and professional category (HCW *vs.* CHW) were added as adjustment variables to the conceptual model to account for the sampling design.

For a SEM model to be identifiable, the number of observations must be greater than the number of parameters to be estimated, i.e. greater than p*(p + 1)/2, where p is the number of variables included. Our model identified 17 variables and 3 adjustment variables, i.e., a minimum of (20*21/2) 210 observations required. We used the weighted least squares mean-variance adjusted (WLSMV on R) estimation method, which allows a relaxation of certain assumptions relating to the distribution of observed variables (normality and linearity) and the use of categorical dependent variables [29]. The model’s goodness-of-fit parameters were verified by a comparative fit index (CFI) > 0.80 and a root-mean-square error of approximation (RMSEA) < 0.05. The validity of these indices enables the model to be evaluated correctly, even when the continuous variables do not meet the normal hypothesis [21,30]. The significance level was set at 5%.

Analyses were performed with RStudio software version 4.1.0, using the “lavaan” function for SEM analysis. As it is based solely on observed variables, the SEM analysis carried out here is a path analysis [20].

## Results

### Description of participants

The sample included 231 respondents (188 HCWs and 43 CHWs), of whom 55.4% were men, with an average age of 37 years (standard deviation (sd)=9) and who had been practicing for an average of 7.4 years (sd=7.1) in the district (Table 2 and detailed results in[16]).

**Table 2.**
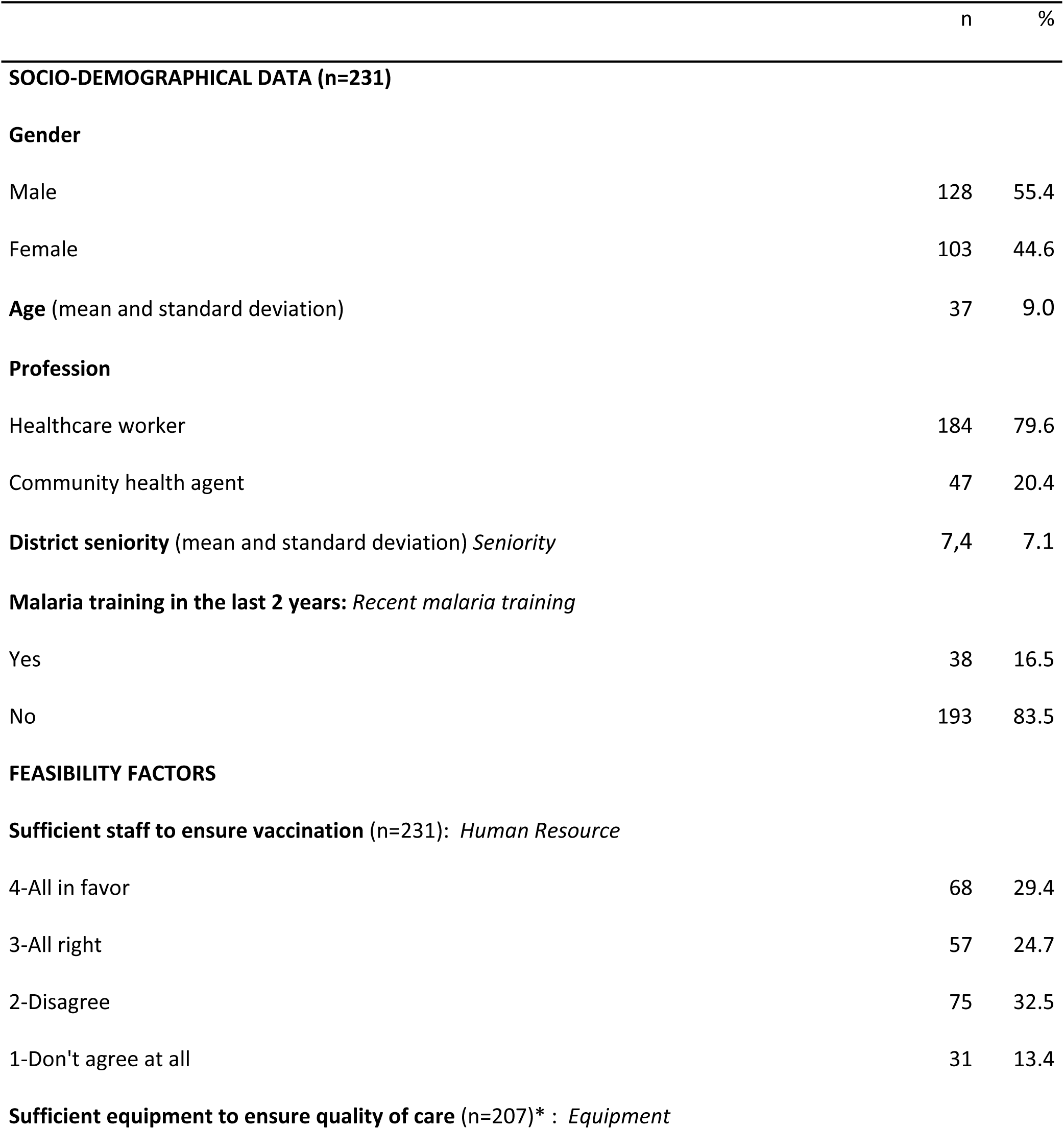
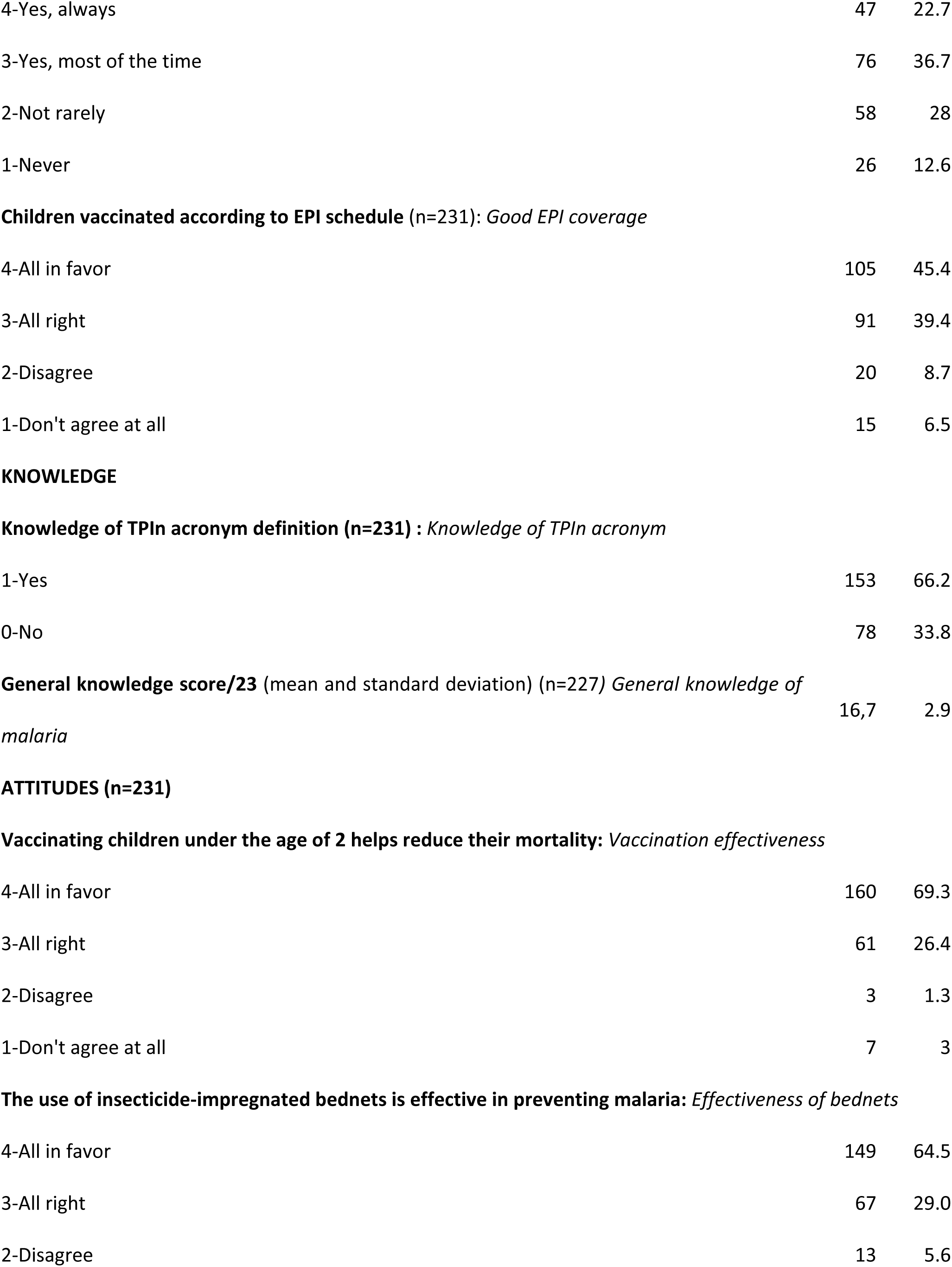
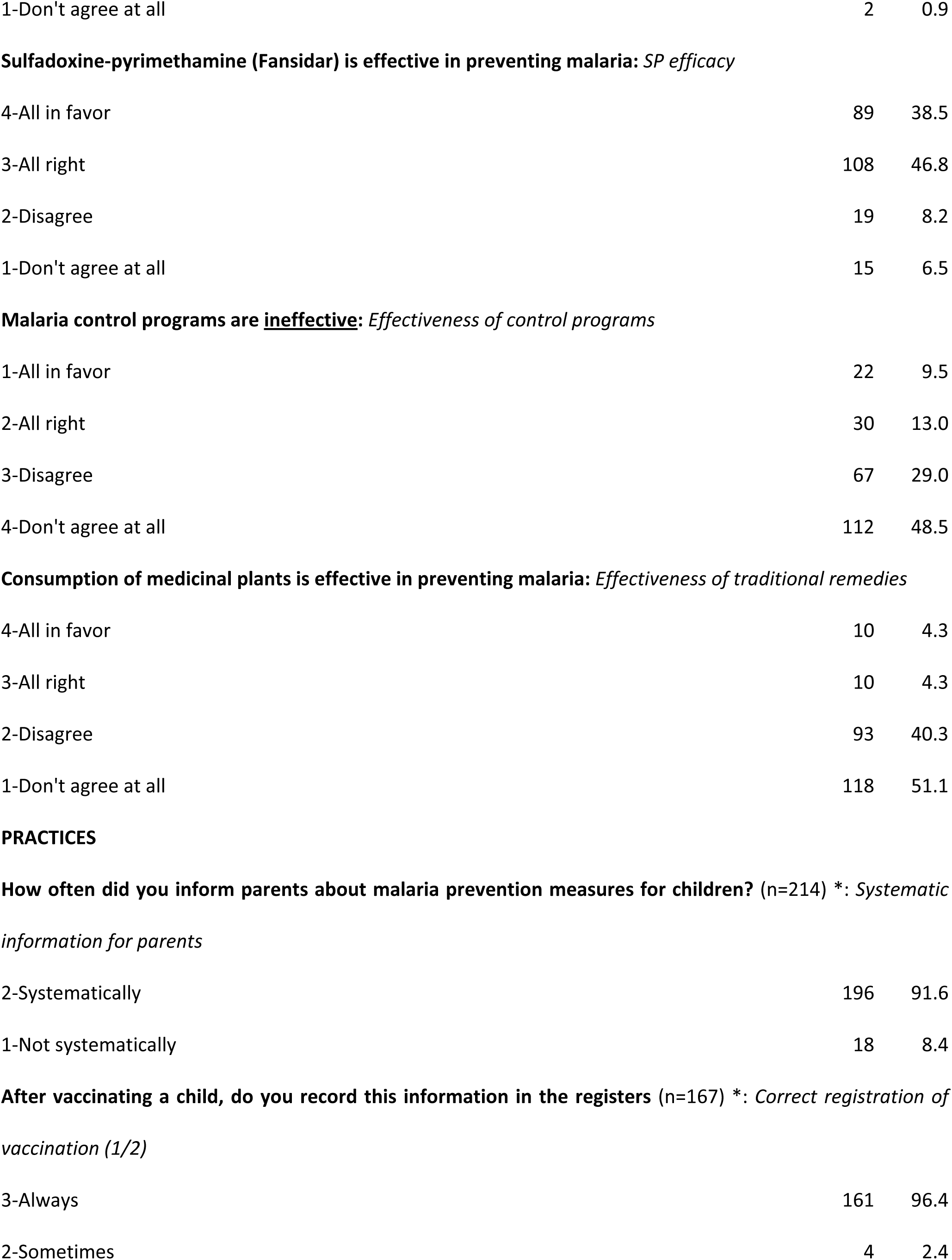
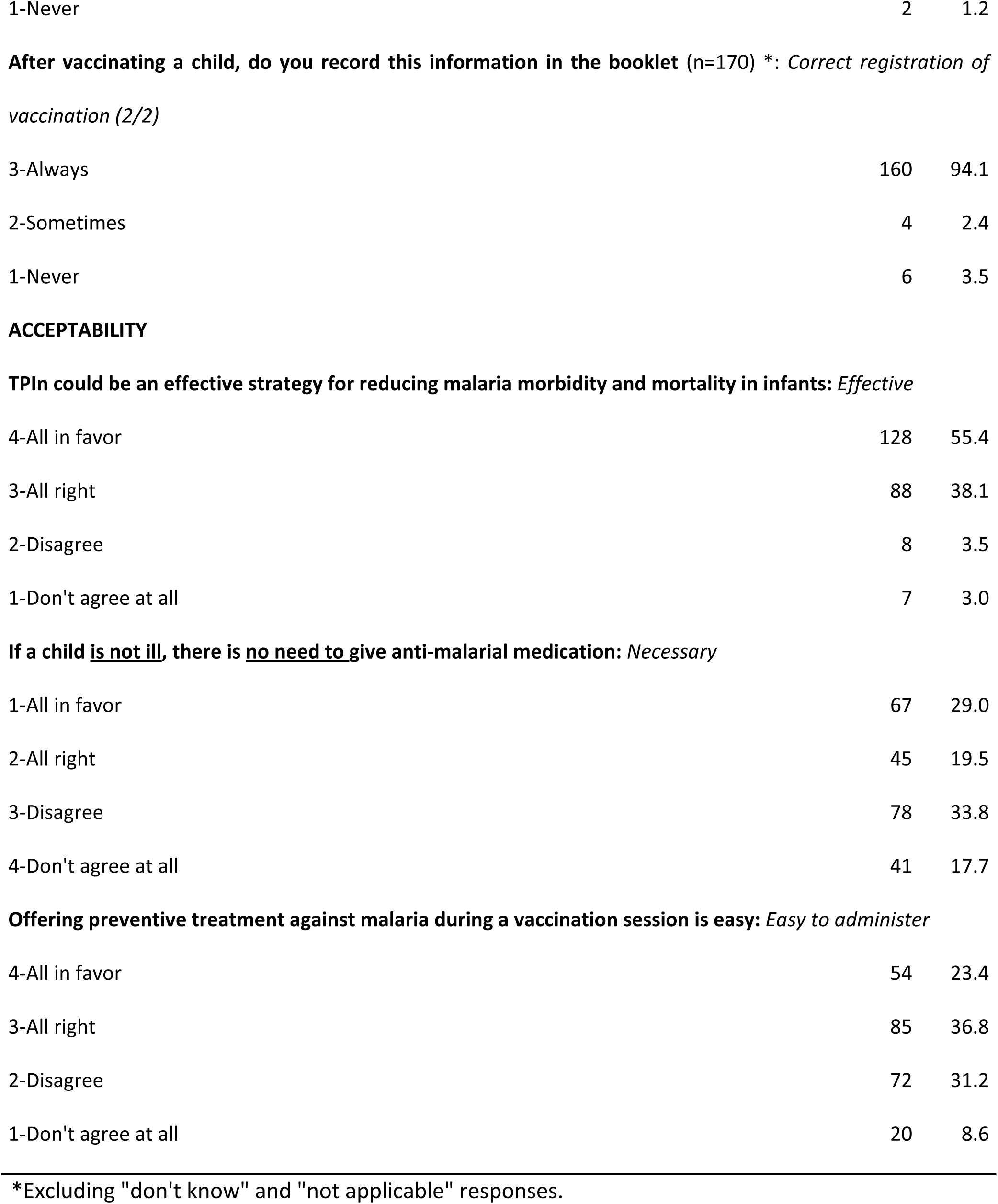
Factors likely to influence providers’ pre-intervention acceptability of the strategy (n=231);

|  | n | % |
| --- | --- | --- |
| **SOCIO-DEMOGRAPHICAL DATA (n=231)** |  |  |
| **Gender** |  |  |
| Male | 128 | 55.4 |
| Female | 103 | 44.6 |
| **Age** (mean and standard deviation) | 37 | 9.0 |
| **Profession** |  |  |
| Healthcare worker | 184 | 79.6 |
| Community health agent | 47 | 20.4 |
| **District seniority** (mean and standard deviation) *Seniority* | 7,4 | 7.1 |
| **Malaria training in the last 2 years: *Recent malaria training*** |  |  |
| Yes | 38 | 16.5 |
| No | 193 | 83.5 |
| **FEASIBILITY FACTORS** |  |  |
| **Sufficient staff to ensure vaccination (n=231): *Human Resource*** |  |  |
| 4-All in favor | 68 | 29.4 |
| 3-All right | 57 | 24.7 |
| 2-Disagree | 75 | 32.5 |
| 1-Don't agree at all | 31 | 13.4 |
| **Sufficient equipment to ensure quality of care (n=207)\* : *Equipment*** |  |  |
| 4-Yes, always | 47 | 22.7 |
| 3-Yes, most of the time | 76 | 36.7 |
| 2-Not rarely | 58 | 28 |
| 1-Never | 26 | 12.6 |
| **Children vaccinated according to EPI schedule (n=231): *Good EPI coverage*** |  |  |
| 4-All in favor | 105 | 45.4 |
| 3-All right | 91 | 39.4 |
| 2-Disagree | 20 | 8.7 |
| 1-Don't agree at all | 15 | 6.5 |
| 1-Yes | 153 | 66.2 |
| 0-No | 78 | 33.8 |
| **General knowledge score/23** (mean and standard deviation) (n=227) *General knowledge of malaria* | 16,7 | 2.9 |
| 4-All in favor | 160 | 69.3 |
| 3-All right | 61 | 26.4 |
| 2-Disagree | 3 | 1.3 |
| 1-Don't agree at all | 7 | 3 |
| **The use of insecticide-impregnated bednets is effective in preventing malaria: *Effectiveness of bednets*** |  |  |
| 4-All in favor | 149 | 64.5 |
| 3-All right | 67 | 29.0 |
| 2-Disagree | 13 | 5.6 |
| 1-Don't agree at all | 2 | 0.9 |
| **Sulfadoxine-pyrimethamine (Fansidar) is effective in preventing malaria: *SP efficacy*** |  |  |
| 4-All in favor | 89 | 38.5 |
| 3-All right | 108 | 46.8 |
| 2-Disagree | 19 | 8.2 |
| 1-Don't agree at all | 15 | 6.5 |
| **Malaria control programs are ineffective: *Effectiveness of control programs*** |  |  |
| 1-All in favor | 22 | 9.5 |
| 2-All right | 30 | 13.0 |
| 3-Disagree | 67 | 29.0 |
| 4-Don't agree at all | 112 | 48.5 |
| **Consumption of medicinal plants is effective in preventing malaria: *Effectiveness of traditional remedies*** |  |  |
| 4-All in favor | 10 | 4.3 |
| 3-All right | 10 | 4.3 |
| 2-Disagree | 93 | 40.3 |
| 1-Don't agree at all | 118 | 51.1 |
| 2-Systematically | 196 | 91.6 |
| 1-Not systematically | 18 | 8.4 |
| 3-Always | 161 | 96.4 |
| 2-Sometimes | 4 | 2.4 |
| 1-Never | 2 | 1.2 |
| 3-Always | 160 | 94.1 |
| 2-Sometimes | 4 | 2.4 |
| 1-Never | 6 | 3.5 |
| **TPIn could be an effective strategy for reducing malaria morbidity and mortality in infants: Effective** |  |  |
| 4-All in favor | 128 | 55.4 |
| 3-All right | 88 | 38.1 |
| 2-Disagree | 8 | 3.5 |
| 1-Don't agree at all | 7 | 3.0 |
| **If a child is not ill, there is no need to give anti-malarial medication: Necessary** |  |  |
| 1-All in favor | 67 | 29.0 |
| 2-All right | 45 | 19.5 |
| 3-Disagree | 78 | 33.8 |
| 4-Don't agree at all | 41 | 17.7 |
| **Offering preventive treatment against malaria during a vaccination session is easy: Easy to administer** |  |  |
| 4-All in favor | 54 | 23.4 |
| 3-All right | 85 | 36.8 |
| 2-Disagree | 72 | 31.2 |
| 1-Don't agree at all | 20 | 8.6 |

\*Excluding "don't know" and "not applicable" responses.

### Model identification and estimation

With 231 observations, the model presented in Fig 1 could be identified and tested, displaying an acceptable fit (CFI=0.82; RSMEA=0.05 [IC95% 0.01-0.07]). Eight hypotheses were validated by the analysis (Table 3).

**Table 3.** SEM analysis results; KAP cross-sectional survey (n=231); MULTIPLY project, Haho district, Togo,

| Assumption | Beta | P-value |
| --- | --- | --- |
| H1 | 0.10 | 0.21 |
| H2 | 0.04 | 0.51 |
| H3* | 0.21 | 0.002 |
| H4* | 0.23 | 0.03 |
| H5* | 0.18 | <0.009 |
| H6 | -0.12 | 0.08 |
| H7 | 0.16 | 0.05 |
| H8* | 0.14 | 0.04 |
| H9 | -0.04 | 0.63 |
| H10* | 0.20 | 0.008 |
| H11* | -0.19 | 0.02 |
| H12* | 0.48 | 0.001 |
| H13 | -0.05 | 0.35 |
| H14 | -0.06 | 0.33 |
| H15* | 0.23 | <0.001 |
| H16 | 0.05 | 0.45 |

\*Assumptions confirmed

### Factors influencing perceived ease of administration of PMC

Over 60% of health care providers perceived that administering PMC during an immunization session would be easy, and almost 55% felt that had sufficient staff to carry out EPI immunization activities (Table 2). The perception of sufficient staff (β= 0.18; p<0.01), good EPI coverage (β= 0.21; p<0.01) and seniority in the district (β= 0.23; p=0.03) were positively and significantly associated with the perception that PMC would be easy to administer (Table 3 and Fig 2).

**Fig 2.**
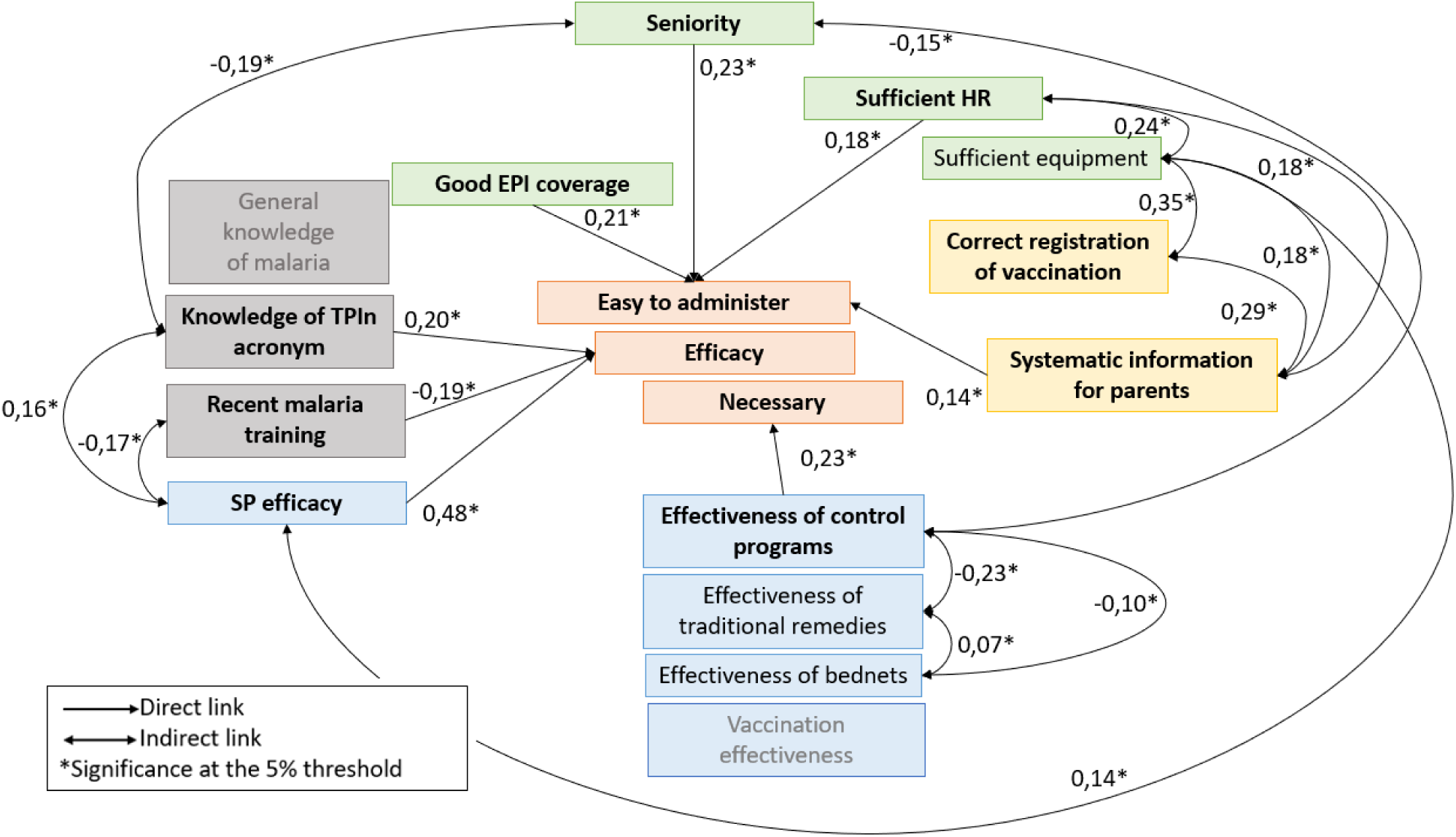
Significant direct and indirect links highlighted by SEM analysis; KAP cross-sectional survey (n=231); MULTIPLY project, Haho district, Togo, November 2021

### Factors influencing the perceived effectiveness of PMC

Most health care providers (93.5%) perceived that PMC would be an effective strategy for reducing malaria in infants (Table 2). Despite high overall knowledge about malaria (mean score 17.3/23), it did not influence the perceived effectiveness of PMC. Factors positively and significantly associated with perceived effectiveness of PMC were knowledge of the IPTi acronym (β= 0.20; p<0.01) and perception of the effectiveness of SP used in prevention (β= 0.48; p<0.01). Having received malaria training in the last 2 years (16.5% of health care providers) was negatively associated with the perceived effectiveness of PMC (β= -0.19; p<0.01). Furthermore, the SEM showed a negative indirect effect between recent malaria training and the perceived efficacy of SP used in prevention (β= -0.17; p=0.04) (Table 3 and Fig 2).

### Factors influencing the perceived need for PMC

Half of the health care providers (51.5%) perceived that there was a need for a malaria preventive treatment to children under 2 years (Table 2). This perception was positively influenced by the perceived the effectiveness of malaria control programs (β= 0.23; p<0.01). The SEM also highlighted a significant negative indirect effect between the perceived effectiveness of malaria control programs and of insecticide-treated bednets (β= -0.10; p=0.04), suggesting that health care providers who perceive bednets as effective are less likely to perceive malaria control programme as effective and to perceive the need for the PMC strategy (Table 3 and Fig 2).

### Links between the 3 components of acceptability

The SEM did not identify direct links between perceived ease of PMC administration, perceived PMC effectiveness, and the need for a malaria preventive treatment, suggesting that these three components of PMC acceptability do not directly influence one another (Table 3). However, the analysis revealed pathways between the various influencing factors, demonstrating the interconnection of the PMC acceptability components. On the one hand, feasibility factors, such as the availability of equipment or staff, not only positively influence the perceived ease of PMC administration but also its perceived effectiveness, as shown by the green pathway in Fig 3. On the other hand, while more experienced providers perceived that PMC would be easy to administer, but did not perceive PMC as necessary or effective, providers with less experience perceived PMC as necessary and effective but not easy to administer, as suggested by the blue pathway in Fig 3.

**Fig 3.**
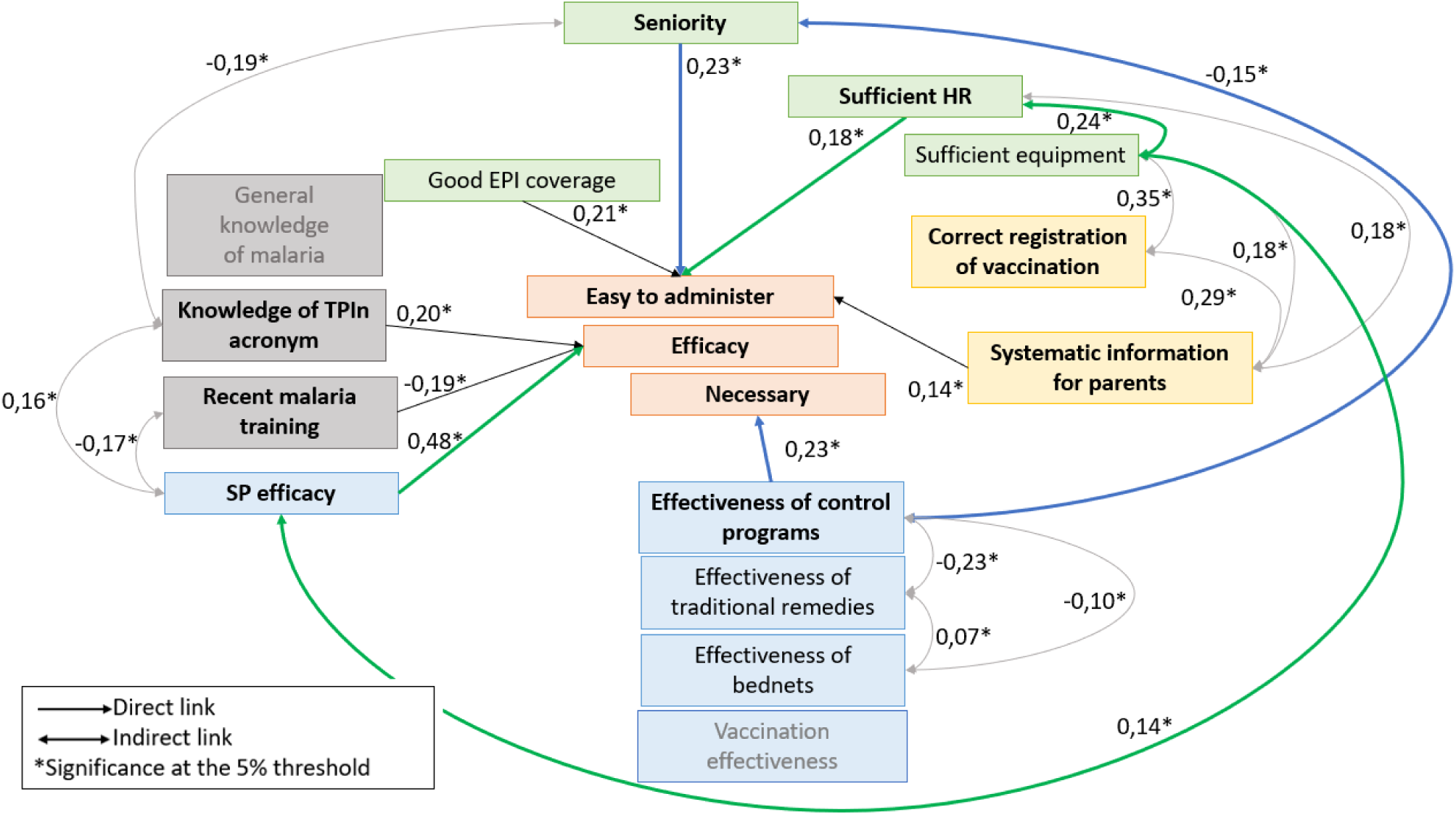
Pathways identified by SEM analysis; KAP cross-sectional survey (n=231); MULTIPLY project, Haho district, Togo, November 2021

## Discussion

The pre-intervention acceptability of PMC was overall high among providers. The SEM analysis highlighted the importance of feasibility factors in shaping the acceptability of PMC, as well as the overall influence of attitudes and, more marginally, of knowledge and practices related to malaria prevention. This in-depth analysis of the multi-faceted concept of acceptability contributed to identify key levers and facilitators for PMC implementation.

### The influence of human resources profile, knowledge and training experience on the acceptability of PMC

According to our results, perceived ease of PMC use was higher among senior providers, while junior providers, trained in chemoprevention and more open to innovation, perceived it as more effective. There is prior evidence of provider seniority having a modulating effect on work motivation [31–33], hence our suggestion overall of building intergenerational teams to enhance collective engagement for the introduction and deployment of PMC strategies. However, further research is needed to better understand the mechanisms underlying this relationship and to determine whether similar patterns would hold across different contexts.

Initial and ongoing training of providers was shown to be crucial to ensure the quality of services [34] and the proper administration of treatments and chemoprevention [9,35]. In our analysis, the completion of a recent training course in malaria was associated with lower perception of SP as an effective preventive measure, and of PMC effectiveness. This result may be explained by the evolution of recommendations concerning the preventive and curative use of SP. Although still recommended for chemoprevention, SP is no longer the sole recommended curative treatment since 2006 [36], due to the increasing resistance of *P. Falciparum* to SP [37]. Reports of therapeutic failures and messages about this resistance may have led to confusion and mistrust in the use of SP in the context of PMC [22,25]. These findings highlight the challenge of aligning training with evolving guidelines and should be interpreted cautiously, as they may reflect broader contextual factors.

In our study, general knowledge of malaria and understanding of the pathology did not influence the acceptability of PMC among health care providers, while knowledge of the specific IPTi acronym influenced its perceived effectiveness. Studies have already shown the link between targeted and technical knowledge and the acceptability of public health interventions [38–41], including PMC [9]. This underlines the need to reinforce ongoing training of the health workforce, prior to PMC implementation, with up-to-date, practical and contextualized scientific content, adapted to realities in the field.

### Combining interventions to combat malaria effectively

In Benin, chemoprevention and vector control strategies (e.g., bednets) were perceived as complementary [23], aligning with international recommendations to combine different malaria control approaches [42]. In our study, the perceived effectiveness of vector control appeared to outweigh that of chemical strategies (mass treatment, seasonal or pregnancy chemoprevention). This divergence suggests that introducing PMC should be accompanied by provider education on the potential synergistic effects of combined interventions on infant malaria morbidity and mortality [42], particularly since the recent inclusion of malaria vaccination in the fight against malaria in children.

### Feasibility, a factor influencing acceptability

Our findings highlight the interactions between the perceived feasibility and the acceptability of PMC. A solid logistical context (positive perceptions about EPI coverage, and human and logistical resources perceived as sufficient) contributed to higher pre-intervention acceptability of the PMC strategy among health care providers. For a pilot intervention, feasibility is fundamental in order to identify practical obstacles, and is a major condition for the acceptability of interventions [43].

### Towards a better understanding of acceptability

According to Batellier, assessing “acceptability” strongly depends on the meaning given to the act of “accepting” and the criteria used during the assessment (attitudes, behaviors, perceptions, etc.) [44]. The results of our SEM analysis confirm that the acceptability of an intervention cannot be reduced to a simple dichotomy (acceptable/not acceptable), but requires a nuanced interpretation that takes into account its complexity (like social or practical aspects) [45] and the diversity of its components [17]. The influence of feasibility factors on the acceptability of PMC among health care providers confirm the complex interdependence between these two concepts [46] and call for further investigation to improve the implementation and effectiveness of health interventions.

Using the TFA to develop the SEM conceptual model contributed to a more in-depth understanding of acceptability theory. Indeed, the development of effective interventions presupposes the identification of the mechanisms by which they induce behavioral change, underlining the importance of integrating theory [47].

### Strengths and limitations

This analysis has several limitations. First, the SEM model and conceptual framework were designed *a posteriori*, based on data from a questionnaire not specifically designed for this type of analysis. Second, the assessment of feasibility relies mainly on perceptions (sufficiency of equipment, personnel), introducing a bias that observational data (number of HR or days out of stock, availability of inputs…) could reduce. Furthermore, we suspect a poor understanding of the question on the need for chemoprophylaxis, due to the wording including a double negative. Not all the components of acceptability identified in TFA [17] have been evaluated and included in this work. Finally, although SEM analysis can identify relationships between variables, it cannot establish causal links.

Nevertheless, the SEM method is particularly well suited to the quantitative analysis of complex concepts such as acceptability, a multifactorial concept with interdependent components[48,49]. Our analysis confirmed that strong logistical support helps to increase the *a priori* acceptability of the PMC strategy, and highlighted several levers for improving the acceptability of the PMC strategy in Togo, including strengthening specific and pragmatic knowledge related to the intervention, providing appropriate training (including on combined approaches to malaria control) and considering the individual experience of providers. These results confirm the findings of a recent literature review showing that knowledge, training and experience are the main factors encouraging providers to comply infection prevention and control measures [38].

## Conclusion

SEM was used to evaluate a pilot intervention combining social science data (KAP data) and statistical modelling. Through research methods based on multidisciplinary approaches, implementation research aims to strengthen health systems to improve the health of populations. Here, the use of SEM has helped to identify the multiple factors influencing the pre-intervention acceptability of a PMC strategy in a rural district of Togo. The use of SEM analyses during pre-intervention studies could help to better understand potential bottlenecks and to develop adaptation strategies to enhance effective implementation of health interventions.

Many African countries have recently included the PMC strategy in their national malaria control plans. However, to date, implementation at national level is only a reality in Sierra Leone, Cameroon and recently in Togo. Our results could help to improve the effectiveness of the introduction, implementation and roll-out of PMC, thereby contributing to the fight against malaria.

## Data Availability

All relevant data are within the manuscript and fully available without restriction.

## Footnotes

1 areas where the prevalence of *P. falciparum* is greater than 10% and/or the annual parasite incidence is greater than 250 cases per 1,000 inhabitants [1]

